# Effects of pharmacogenomic (PGx) testing on clinical pain management prescriptions, a retrospective study

**DOI:** 10.1101/2021.06.21.21258931

**Authors:** Christian Tagwerker, Mary Jane Carias-Marines, David J. Smith

## Abstract

Current deficits in effectively utilizing PGx testing in clinical practice include limited awareness and training of healthcare professionals, routine ordering of assays investigating up to 5 genes and lack of concise reporting of dosing guidelines and drug-drug-interactions. A novel deep sequencing (>1000X) PGx panel is described encompassing 23 genes and 141 SNPs or indels combined with PGx dosing guidance, drug-gene-interaction (DGI) and drug-drug-interaction (DDI) reporting to prevent adverse drug reaction events. During a 2-year period, patients (n = 171) were monitored in a pain management clinic. Urine toxicology, PGx reports, and progress notes were studied retrospectively for changes in prescription regimens before and after the PGx report was made available to the provider.

Among patient PGx reports with medication lists provided (n = 146) 57.5% showed one or more moderate and 5.5% at least one serious pharmacogenetic interaction. 66% of patients showed at least one moderate and 15% one or more serious drug-gene or drug-drug-interaction. A significant number of active changes in prescriptions based on the PGx reports provided was observed for 85 patients (83%) for which a specific drug was either discontinued, switched within the defined drug classes of the report or a new drug added.

Preventative action was observed for all serious interactions and only moderate interactions were tolerated for lack of other alternatives. This study demonstrates a successful implementation of PGx testing utilizing an extended PGx panel combined with a customized, informational report to help improve clinical outcomes.

## 1 Introduction

Over the last decade, there has been considerable growth in the use of pharmacogenomic (PGx) testing due to increased awareness of patients developing moderate to serious adverse drug reaction reactions (ADRs) attributed to individual genetic variation. The US Food and Drug Administration (FDA) genetic testing recommendations and black box warnings for 262 individual drug labels are covered by 86 genetic biomarkers (status December 2018) relating to dosage and administration, warnings, precautions, drug interactions, adverse reactions or clinical pharmacology [1]. For example, codeine, a frequently prescribed opiate present in TYLENOL #3^®^ (acetaminophen with codeine), contains the boxed warning: “*Death Related to Ultra-Rapid Metabolism of Codeine to Morphine. Respiratory depression and death have occurred in children who received codeine in the post-operative period following tonsillectomy and/or adenoidectomy and had evidence of being ultra-rapid metabolizers of codeine. Deaths have also occurred in nursing infants who were exposed to high levels of morphine in breast milk because their mothers were ultra-rapid metabolizers of codeine*.” A survey involving clinicians from academic medical centers showed 99% agreed that PGx variants would influence a patients’ response to drug therapy and should be acted upon when a clinically significant drug-genome interaction was present (92%) [2]. Previous studies have shown that over 80% of patients can carry at least one functional gene variant influencing one of the 100 most prescribed medications in the US and the rate of rehospitalization can be significantly reduced by implementation of PGx test recommendations [3]–[7].

Recommendations for actionable prescribing decisions are routinely based on clearly defined, peer-reviewed guidelines with different evidence levels (Levels 1-4) issued by international pharmacogenetic consortia and professional societies such as the Clinical Pharmacogenetics Implementation Consortium (CPIC) and maintained in high-quality public and expert-curated databases, including PharmVar and PharmGKB [8]–[11]. Currently most laboratories conducting PGx testing use targeted genotyping technologies to screen for specific variants to determine adverse drug reactions. Examples of these technologies include single or multiplexed PCR assays combined with Taqman hydrolysis probe chemistry, microarrays (ThermoFisher Scientific), mass spectrometry (Agena Biosciences), bead-based immunoassays (Luminex) or Next-Generation Sequencing (NGS) assays (Illumina) [12]–[14]. This study describes application of PGx report recommendations (including genetic based dosing guidance (PGx), drug-gene interaction (DGI) and drug-drug interaction (DDI) based guidance) compared to quantitative urine drug toxicology (UDT) reports. UDT reports were evaluated in a pain management setting before and after application of PGx panels encompassing 23 genes to prevent adverse drug reaction events (**Fig 1**).

**Fig 1.**
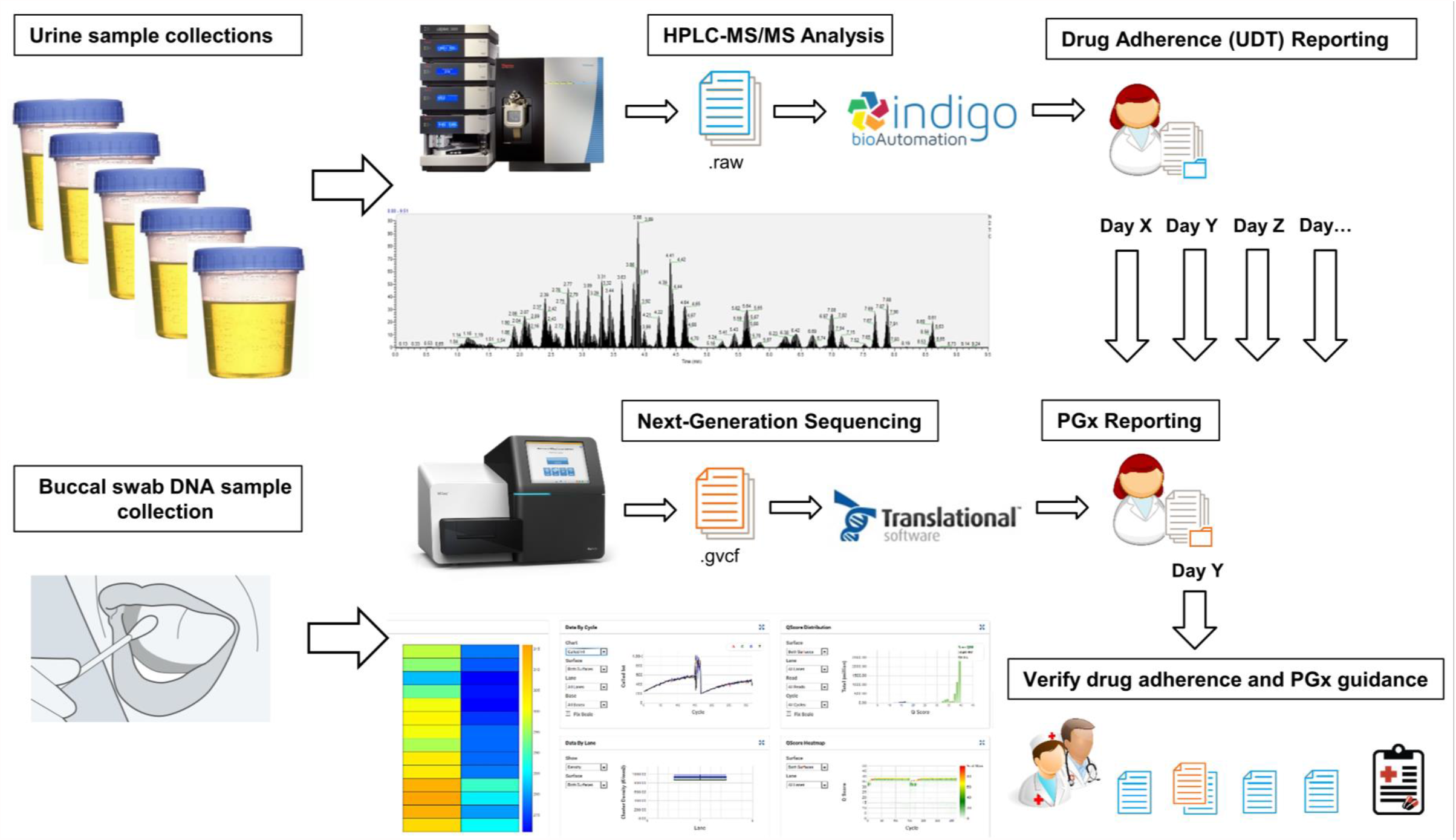
Overview of this study to determine implementation of PGx report recommendations as compared to urine drug adherence reports in a pain management setting after application of a deep sequencing PGx panel.

## 2 Materials and Methods

This study was conducted in accordance with the Declaration of Helsinki with written informed consent from each patient. Patient data collection and summaries were approved by the Alcala Pharmaceutical Inc. Institutional Review Board (IORG0010127, IRB00012026, #R003). All test samples derived from human subjects were de-identified of their health information as defined by HIPAA guidelines. Patient data for comparison of urine drug adherence testing before and after PGx reporting were obtained retrospectively from patients (n = 171) in a pain management clinic representing an ethnically diverse patient population from 2016 to 2018 within the western United States. Available data includes de-identified pre- and post-PGx medication lists, PGx and urine drug adherence data.

### 2.1 Genes

23 genes were selected based on having the most clinical utility in PGx at the time of design in April 2016 (*ADRA2A, CES1, COMT, CYP1A2, CYP2C19, CYP2C9, CYP2D6, CYP3A4, CYP3A5, DRD1, DRD2, F2, F5, GNB3, HTR1A, HTR2A, HTR2C, MTHFR, OPRM1, SLC6A2, SCL6A4, SLCO1B1, VKORC1*).

### 2.2 Selection of Target Regions

Online probe design was performed by entering target regions into Design Studio (DS) software (Illumina) [15]. 75 target regions were covered by 82 amplicons with an average amplicon size of 250 basepairs (bp). Possible gaps in target coverages, repeats and GC-rich regions that could interfere with optimal amplification of all desired regions were identified in 3 iterations (design 32844, 32865 and 98659) and optimized for TruSeq Custom Amplicon Low Input (TSCA-LI) assay technology (Homo sapiens (UCSC hg19), Variant source: 1000 Genomes). Predicted coverage of the full region of interest was 100% with all amplicons showing scores at 100%. Unique reference single-nucleotide polymorphism cluster ID (rsID) numbers were assigned per target coordinate and region (**Supplementary Table 1**). Oligonucleotide probes were synthesized and pooled at Illumina (San Diego, CA) into a Custom Amplicon Tube (CAT).

### 2.3 DNA Isolation and Genotyping

Genomic DNA (gDNA) was isolated from up to 4 buccal swab specimens provided by the pain management clinic using PureLink Genomic DNA Isolation (ThermoFisher Scientific, Carlsbad, CA) and Agencourt DNAdvance Genomic DNA Isolation kits (Beckman Coulter, Indianapolis, IN). Quality and concentration of gDNA were determined using Qubit 3.0 Fluorometric Quantitation (ThermoFisher Scientific). Next-Generation Sequencing (NGS) was carried out on a MiSeq system (Illumina, San Diego, CA) with 2×150 bp paired-end reads using the TruSeq® Custom Amplicon v1.5 Targeted Resequencing workflow (Illumina) for up to 24 samples per plate. HYB and EXT_LIG programs were as described in the TSCA-LI protocol. Amplification was carried out at 32 cycles (<96 amplicon plexity). After cleanup and normalization by AMPure XP magnetic beads, pooled libraries were denatured at 98°C for 2 minutes and cooled on ice for 5 minutes. Denatured PhiX control (12.5 pmol/L) was spiked into the library pool at 1% and loaded onto an Illumina MiSeq instrument at 7 pmol/L for automated cluster generation and sequencing according to the manufacturer’s instructions. All targets and 50 bp flanking regions were sequenced, the capture region totaled approximately 20 kb.

### 2.4 Data Analysis and Interpretation

The TruSeq Amplicon workflow version 1.0.0.61 on the MiSeq instrument was used to perform primary analysis by Real Time Analysis (RTA, version 1.18.54) during the sequencing run. Base calls of indexed raw sequence reads and demultiplexing were performed using bcl2fastq. MiSeq Reporter version 2.6.2.3 performed secondary analysis on base calls and quality scores generated on-instrument by the RTA software and evaluated short regions of amplified DNA for variants. Clusters from each sample were aligned against amplicon sequences from the provided manifest file (Design 98659). The first read was evaluated against the probe sequence for each amplicon in the manifest, which is the reverse complement of the DLSO (Downstream Locus-Specific Oligo). If the start of the read matches (with at most 1 mismatch) a probe sequence, the read was aligned against the target(s) for that probe sequence. If no such match was found for the read, MiSeq Reporter checked for any probe sequence which is matched with fewer than six mismatches, and attempted to align against these amplicons. For paired-end data, the second read was handled similarly, except that read 2 is compared to ULSO (Upstream Locus-Specific Oligo) sequences. After the probe sequence (ULSO or DLSO) was matched, adapter sequences were removed and trimmed reads mapped to the human reference genome (GRCh37 hg19) using banded Smith-Waterman alignment generated in .bam file format. Maximum indel length is normally 10 bp, but was overridden using the sample sheet setting CustomAmpliconAlignerMaxIndelSize set to 250 (higher values improve indel sensitivity but impact workflow speed). Other sample sheet settings included IndelRepeatFilterCutoff set to 1, MinimumCoverageDepth = 1, VariantMinimumGQCutoff = 1, VariantFilterQualityCutoff = 1, VariantCaller = GATK, VariantAnnotation = MARS, outputgenomevcf = TRUE. Genome Analysis Toolkit (GATK, Broad Institute) identifies variants, and writes .vcf and .gvcf output files to the Alignment folder. SNPs and short indels were identified using GATK for each sample and false discovery rates for each variant were evaluated using coverage (read depth), the Qscore (quality) and GQX value (a conservative measure of genotype quality derived from the minimum of the GQ and QUAL values listed in the .vcf file). The Qscore predicts probability of an erroneous base call (Q20 represents the probability to call an erroneous base out of 100, reflecting an accuracy of the sequenced base at 99%, Q30 = 99.9%, Q40 = 99.99%, etc.). Coverage for a defined region is the total number of reads passing quality filters at this position representing a given nucleotide. Only variants showing Qscores and GQX values >30 and coverage >100X were considered in this study. Two positive genomic DNA controls (PC1 and Coriell cell line NA19920 gDNA) and one negative (RS1 buffer) control were sequenced per plate (up to 48 samples). All 167 mutation sites covering 141 SNPs, 2 gender probes and one indel (43-44 bp insertion in the *SLC6A4* promoter region – short (S) or long form (L) – see Supplementary Table S1) within the 23 genes identified by MiSeq Reporter were reviewed for each sample in VariantStudio software (Illumina) assisted by the PASS filter function. Gender probes were matched to the provided gender in the sample requisition. All samples and positive controls were imported as .gvcf files into a customized portal through Translational Software™ Inc. (TSI, Bellevue, WA) [16]. After entry of *SLC6A4* indel S/S, La/La, La/Lg or Lg/Lg variants and *CYP2D6* deletion or duplication data transfer of all variants and phenotype calls were reviewed for samples and quality controls prior to custom medical report generation for each patient. Translational Software provides interpretations of specific variants for dosing guidance and drug-drug interaction (DDI) warnings provided by a third-party agreement with First Data Bank (FDB). (**Supplementary Table 2**). Control genomic DNA from NA18861, NA18868, NA19920, NA19226 purchased from Coriell Cell Biorepositories and internal positive controls were used for validation of the TSCA-LI workflow with design 98659, CNV/Indel assay validations and for evaluation of the data interpretation software by TSI.

### 2.5 Copy Number Variation and Indel Assays

Copy number variations (CNVs) of *CYP2D6* were identified with two different PCR reactions for detection of *CYP2D6*\*XN duplication or *CYP2D6*\*5 deletion events by long-range PCR as previously described [17], [18]. Ten nanograms of input gDNA was used with Takara LA *Taq* polymerase (Takara Bio USA, San Diego, CA) carried out according to the manufacturer’s instructions. The long-range PCR conditions for duplication testing were as follows: initiation at 94°C for 2 min, 27 cycles of 98°C for 20 sec, 61.4°C for 20 sec and 68°C for 10 min, and termination at 72°C for 10 min. PCR conditions for deletion tests were the same except annealing was at 65°C for 25 sec and extension at 68°C for 5 minutes with 25 cycles and termination at 72°C for 6 min. Long-range PCR products were analyzed by 1.0% agarose gel electrophoresis. The presence of a 10 kB fragment (by primers CY_DUP_5 and CY_DUP_3) indicated duplicated or multicopy *CYP2D6* alleles and a 3.5 kb product (by primers CY_DEL_5 and CY_DEL_3) was indicative of the deletion (*CYP2D6*\*5 allele). Amplification of the short (S) and long (L) variant of the 5-HTT gene-linked polymorphic region (5-HTTLPR) of SLC6A4 was accomplished with oligonucleotide 5-HTTF, corresponding to nucleotide positions −1346 to −1324 and 5-HTTR (positions from −910 to −888) as previously described [19], [20], except amplification was performed in 25 μl containing 10 ng of genomic DNA, 1.5 mM MgCl_2_, 200 μM dNTPs, 1X Colorless GoTaq® Flexi buffer, 0.4 μM of each primer and 1 U of hot start GoTaq DNA polymerase (Promega Biosciences, San Luis Obispo, CA). Initial denaturation was performed at 98°C for 3 min, followed by 35 cycles at 94°C for 1 min, 64°C for 30 sec and 72°C for 2 min. PCR products were resolved by 2% agarose gel electrophoresis. 458 and 415 bp fragments indicated the L/S genotype for SLC6A4, single 415 bp bands or 458 bp bands (no double band profile) indicated the S/S and L/L genotypes, respectively. All primer sequences are listed in **Supplementary Table 3**.

### 2.6 Drug Adherence Testing

All PGx reports were compared to urine toxicology reports generated before or after clinicians received the PGX report. Urine toxicology reports reviewed by clinical laboratory scientists with ASCENT™ review software (IndigoBio Automation, Carmel, IN) [21] were made available by routine HPLC-MS/MS presumptive and confirmatory urine drug testing at ATAS from 2016-2018 [22].

## 3 Results and Discussion

Analytical sensitivity (call rate) was determined at >97.1% by positive agreement of all 141 variants including sex determination through 2 SRY gender probes and CNVs/Indels. Genomic DNA ranging from 0.64–26 ng/µL (5-195 ng input gDNA) was sequenced across three validation plate runs with 68 positive control samples showing unambiguous genotypes. Buccal swabs were stored for up to 14 days at 4°C prior to gDNA preparation, gDNA storage stability at 4°C was confirmed for up to 6 days and up to 6.5 months for storage at -20°C with up to 10 freeze/thaw cycles to yield high quality (> 99.3%) genotyping results **Supplementary Table 4**.

All alleles covered per gene target(s) and resulting phenotypes were routinely described in the test details section in each PGx report (**Table 1**) following the results for pharmacogenetic (PGx) based dosing and drug-gene (DGI) or drug-drug interactions (DDI) (**Fig 2**).

**Table 1:** Example of PGx report test details summary and alleles covered.

| Test Details |  |  |  |
| --- | --- | --- | --- |
| Gene | Genotype | Phenotype | Alleles Tested |
| *CYP2C9* | *1/*1 | Normal Metabolizer | *2, *3, *4, *5, *6, *7, *8, *11, *14, *27 |
| *CYP2C19* | *2/*17 | Intermediate Metabolizer | *2, *25, *3, *4, *4B, *5, *6, *7, *8, *9, *10, *12, *14, *15, *17 |
| *CYP2D6* | *1/*2 | Normal Metabolizer | *2, *3, *31, *33, *4, *4M, *46, *49, *53, *6, *7, *8, *9, *10, *11, *12, *14A, *14B, *15, *17, *29, *35, *38, *41, *44, *5 (gene deletion), XN (gene duplication) |
| *CYP3A5* | *3/*3 | Poor Metabolizer | *1D, *2, *3, *3B, *3C, *4, *6, *7, *8, *9 |
| *CYP3A4* | *1/*1 | Normal Metabolizer | *2, *4, *5, *8, *11, *12, *13, *16A, *16B, *17, *18A, *18B, *20, *22 |
| *VKORC1* | -1639G>A A/A | High Warfarin Sensitivity | -1639G>A, 1542G>C, 5808T>G, 1173C>T, rs11540137, rs13337470, 698C>T, 2255C>T, 3730G>A |
| *CYP1A2* | *1F/*1F | Normal Metabolizer – Higher Inducibility | *1C, *1D, *1E, *1F, *1J, *1K, *1L, *1V, *1W, *7 |
| *SLCO1B1* | 521T>C T/C | Decreased Function | 388A>G, 521T>C, 467A>G, -11187G>A, 1865+248G>A |
| *COMT* | Val158Met A/G | Intermediate COMT Activity | Val158Met |
| *OPRM1* | A118G A/A | Normal OPRM1 Function | A118G |
| *HTR2C* | -759C>T C/T | Heterozygous for the C Allele (rs3813929) | -759C>T, 2565G>C |
| *SLC6A4* | S/La | Decreased Serotonin Transporter Expression | La, S, Lg |
| *ADRA2A* | C-1291G C/C | Homozygous for C Allele | C-1291G |
| *SLC6A4* | 463T>G A/A | Homozygous for A Allele | La, S, Lg |
| *HTR2A* | rs7997012 G/G | Homozygous for G Allele (rs7997012) | 102C>T, -1483G>A, rs7997012 |
| *HTR2C* | 2565G>C C/C | Homozygous for C Allele (rs1414334) | -759C>T, 2565G>C |
| *HTR2A* | -1438G>A, T/T | Homozygous for T Allele (rs6311) | 102C>T, -1483G>A, rs7997012 |
| *DRD2* | -241A>G, T/C | Heterozygous for rs1799978 C Allele | -241A>G, rs2283265, 939T>C, 957C>T |
| *DRD2* | rs2283265 C/C | Homozygous for rs2283265 C Allele | -241A>G, rs2283265, 939T>C, 957C>T |
| *MTHFR* | 1298A>C AA<br>677C>T CC | No Increased Risk of Hyperhomocysteinemia | 677C>T, 1298A>C, 1305C>T |
| *MTHFR* | 677C>T CC | Normal MTHFR Activity | 677C>T, 1298A>C, 1305C>T |
| Factor II<br>Factor V Leiden | 20210G>A GG<br>1691G>A GG | No increased Risk of Thrombosis | 20210G>A, 1691G>A |

**Fig 2.**
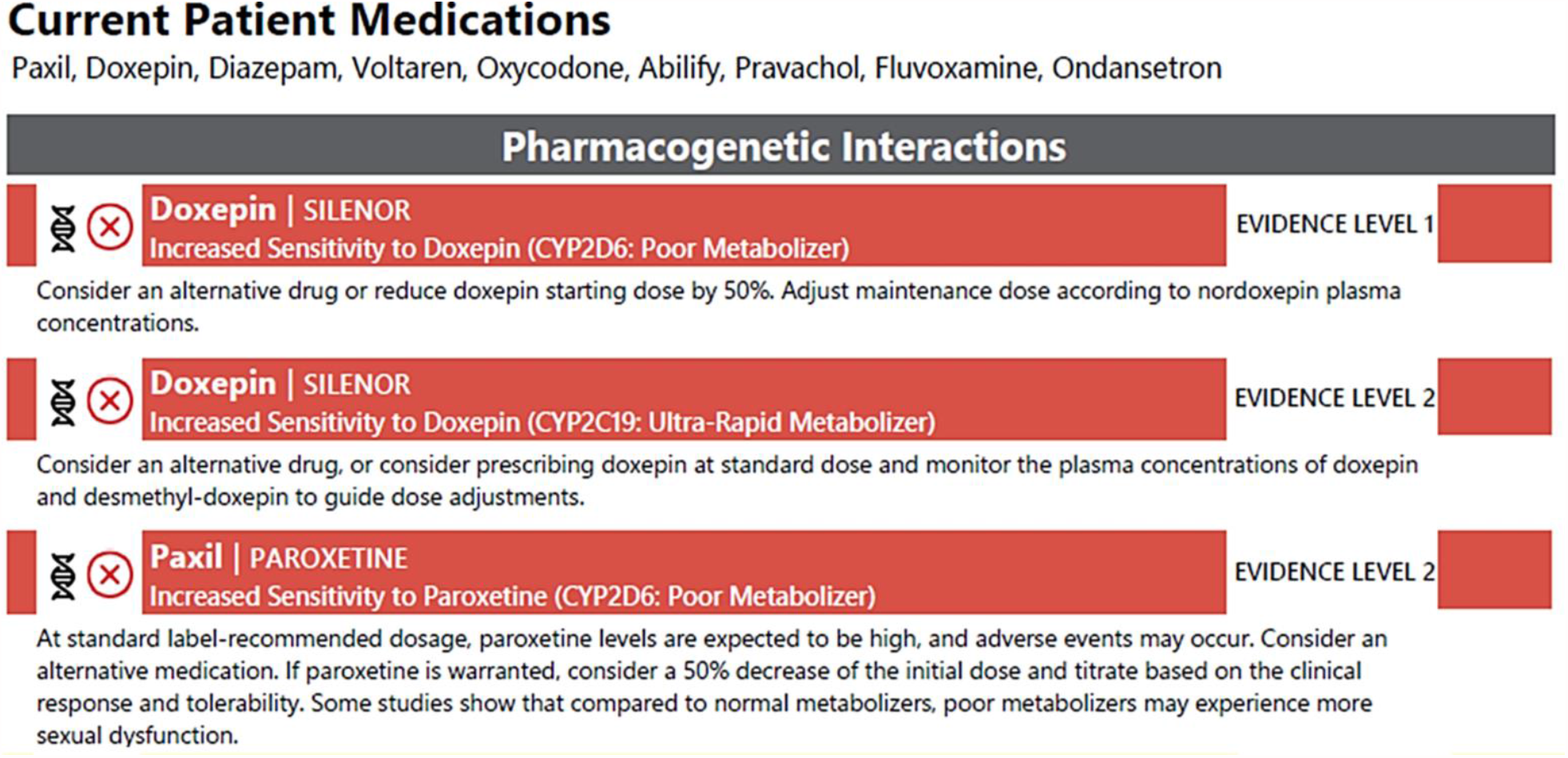
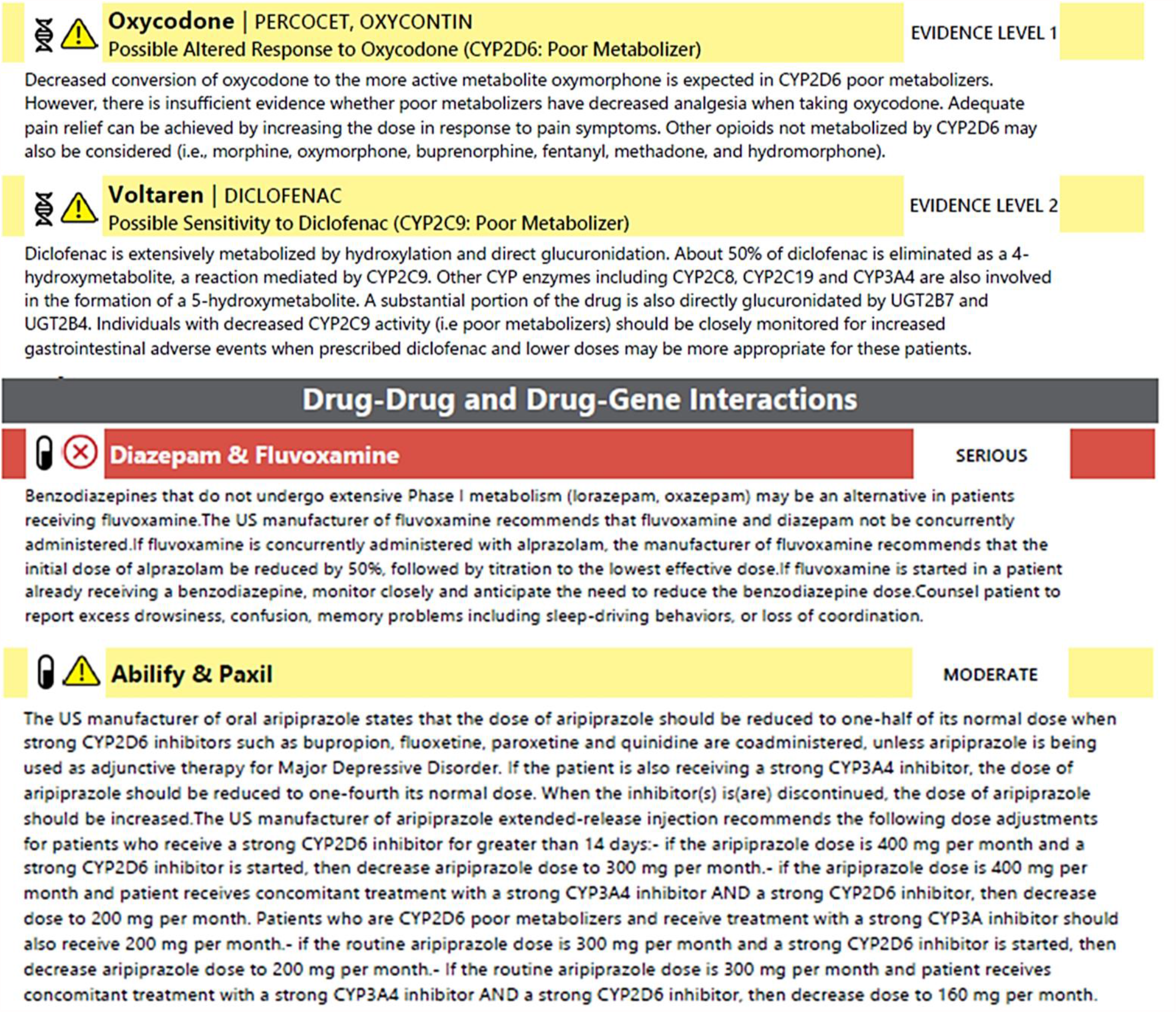
Example of PGx report results showing PGx dosing guidance, drug-gene (DGI) and drug-drug interactions (DDI). Evidence Level 1 descriptions were actionable with established, evidence-based clinical guidelines issued by international pharmacogenetic consortia, professional societies or regulatory bodies (CPIC, DPWG, FDA, EMA, CPNDA, ACMG). Evidence Level 2 descriptions were informative, requiring further investigations. PGx dosing guidance, drug-gene (DGI) and drug-drug interactions (DDI) were further marked as either yellow (MODERATE) or red (SERIOUS) interactions.

Phenotypes and associated genotypes were summarized in **Table 2** with an overview of population frequencies compared to this cohort.

**Table 2:**
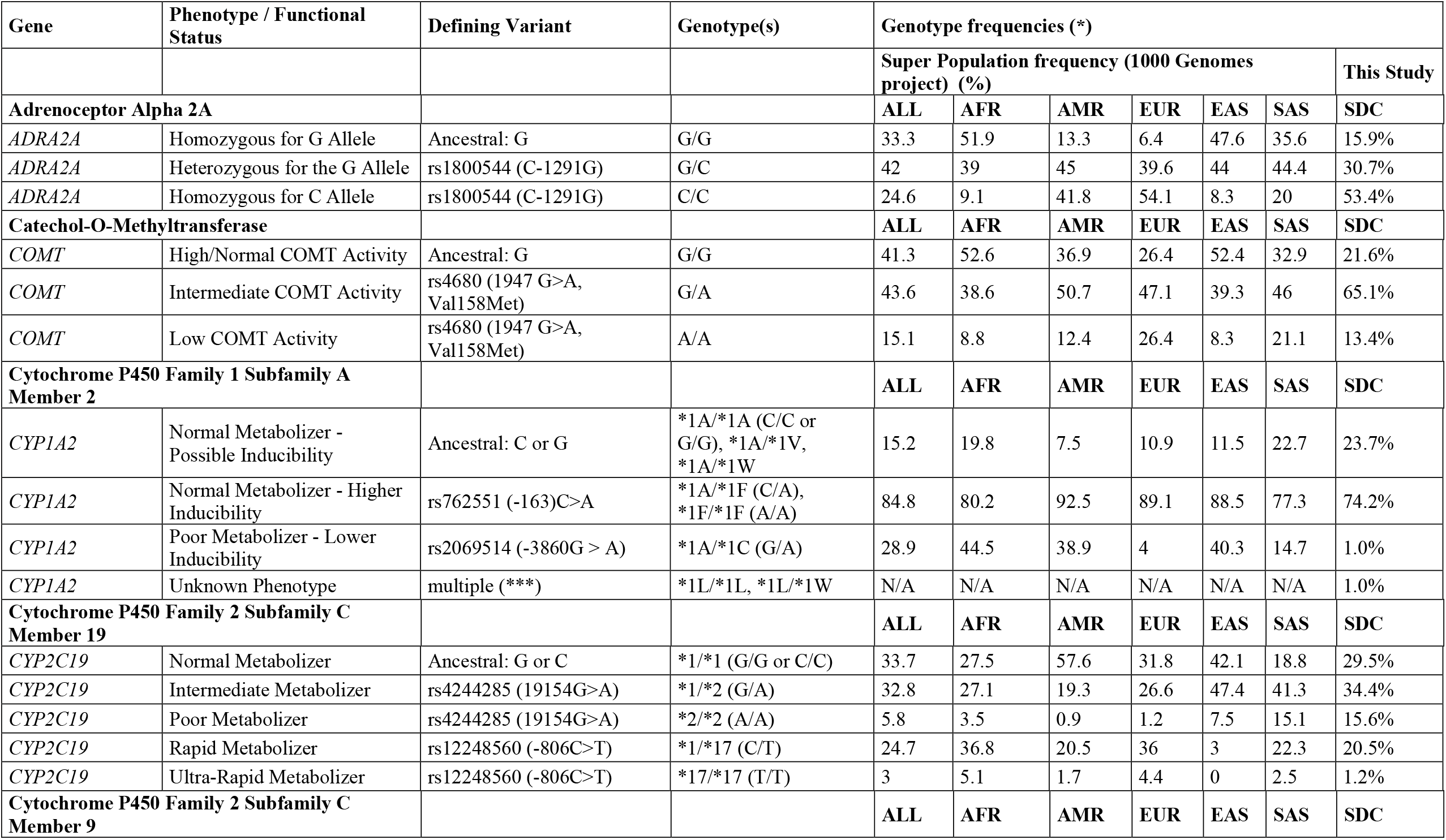
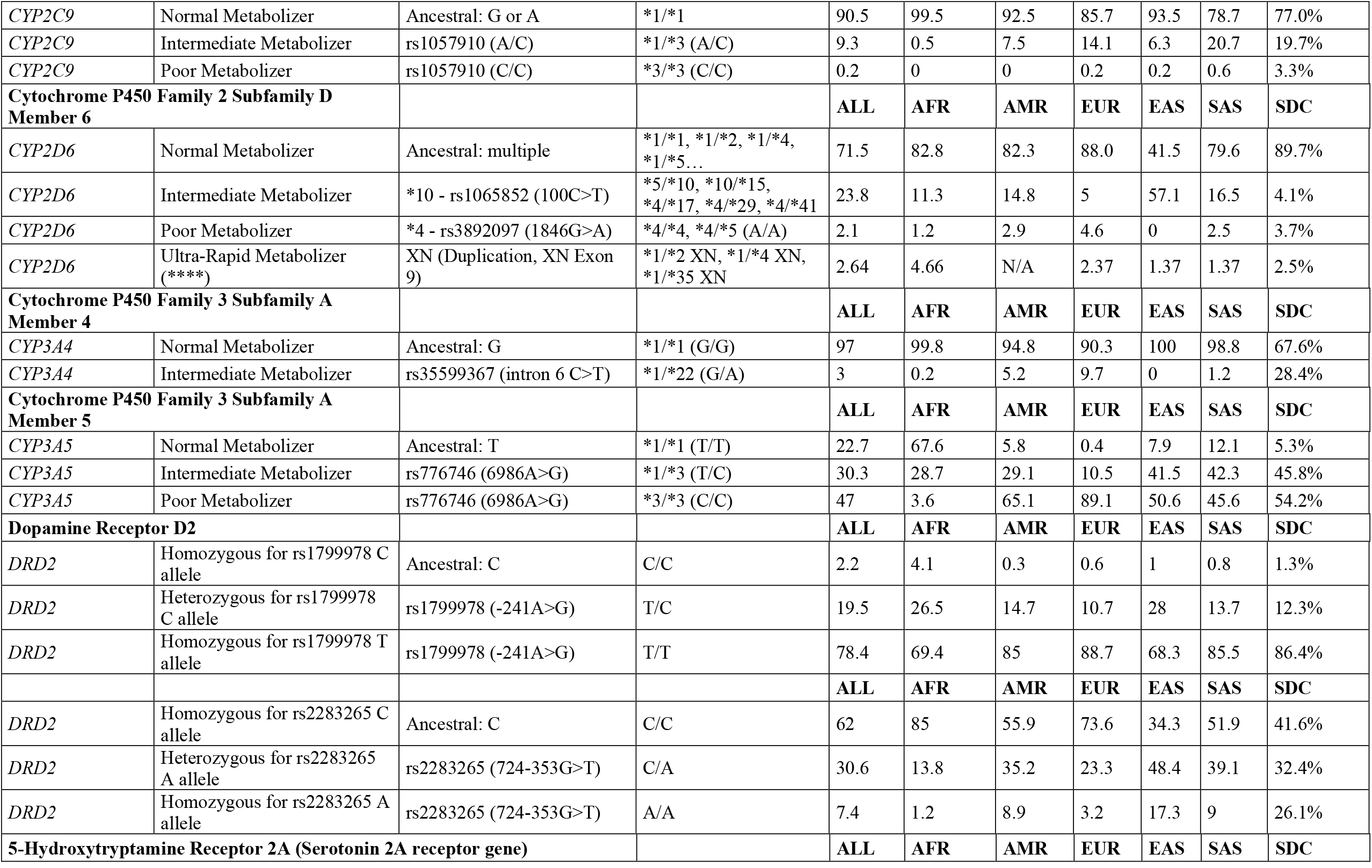
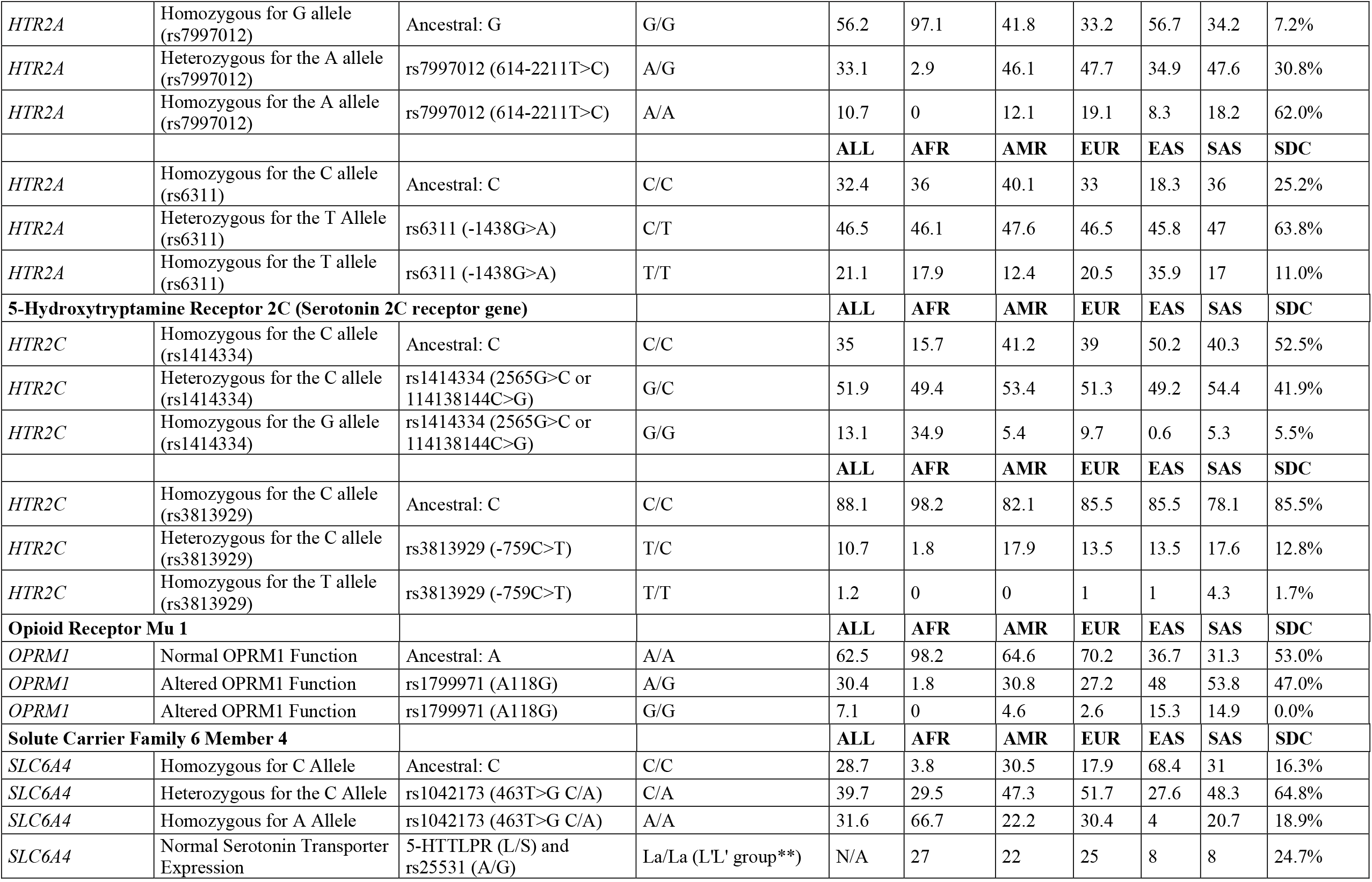
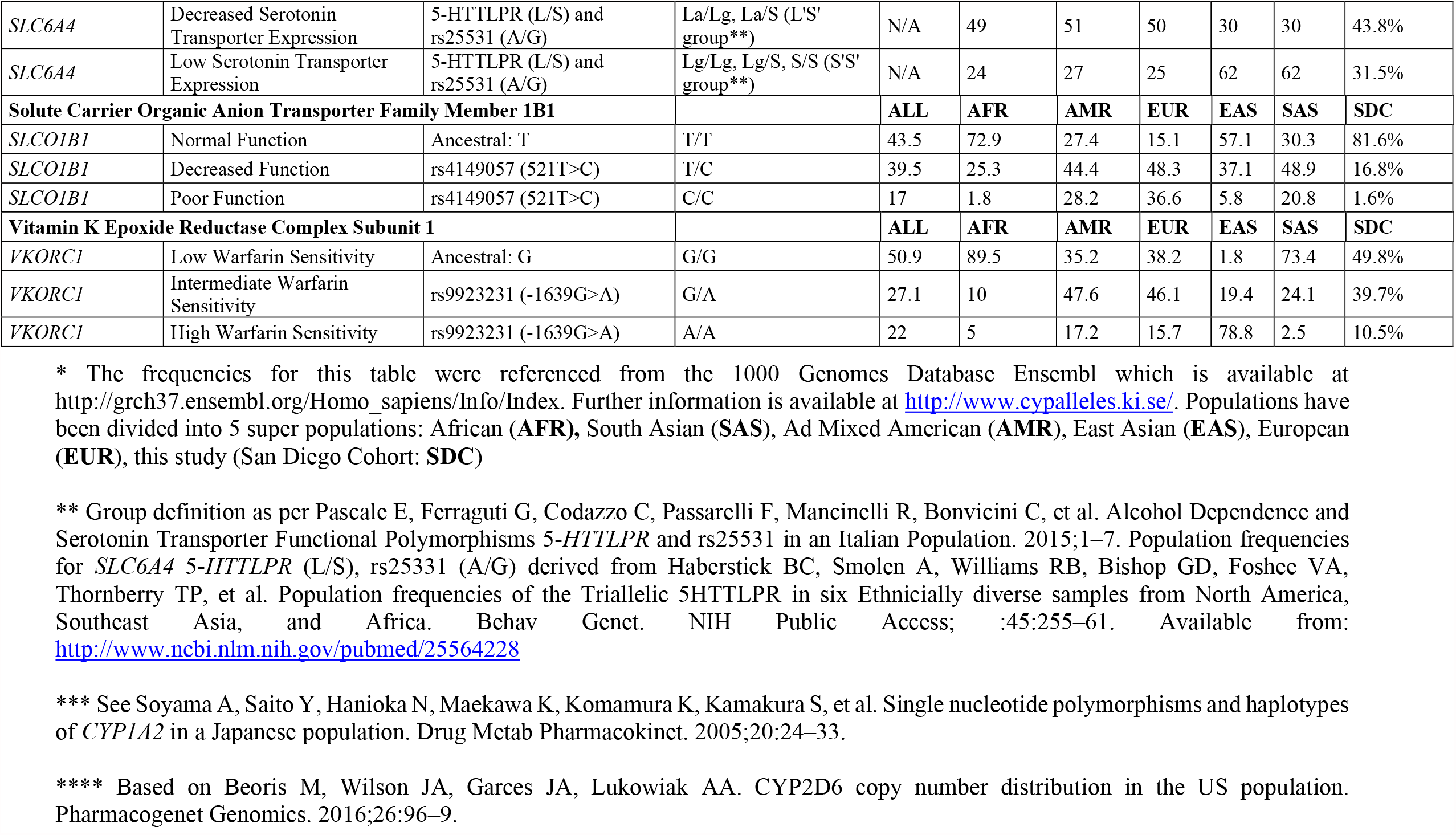
Observed phenotypes and associated genotypes with an overview of population frequencies compared to this study (n = 171).

| Gene | Phenotype / Functional Status | Defining Variant | Genotype(s) | Genotype frequencies (*) |  |  |  |  |  |  |
| --- | --- | --- | --- | --- | --- | --- | --- | --- | --- | --- |
|  |  |  |  | Super Population frequency (1000 Genomes project) (%) |  |  |  |  |  | This Study |
| Adrenoceptor Alpha 2A |  |  |  | ALL | AFR | AMR | EUR | EAS | SAS | SDC |
| ADRA2A | Homozygous for G Allele | Ancestral: G | G/G | 33.3 | 51.9 | 13.3 | 6.4 | 47.6 | 35.6 | 15.9% |
| ADRA2A | Heterozygous for the G Allele | rs1800544 (C-1291G) | G/C | 42 | 39 | 45 | 39.6 | 44 | 44.4 | 30.7% |
| ADRA2A | Homozygous for C Allele | rs1800544 (C-1291G) | C/C | 24.6 | 9.1 | 41.8 | 54.1 | 8.3 | 20 | 53.4% |
| Catechol-O-Methyltransferase |  |  |  | ALL | AFR | AMR | EUR | EAS | SAS | SDC |
| COMT | High/Normal COMT Activity | Ancestral: G | G/G | 41.3 | 52.6 | 36.9 | 26.4 | 52.4 | 32.9 | 21.6% |
| COMT | Intermediate COMT Activity | rs4680 (1947 G>A, Val158Met) | G/A | 43.6 | 38.6 | 50.7 | 47.1 | 39.3 | 46 | 65.1% |
| COMT | Low COMT Activity | rs4680 (1947 G>A, Val158Met) | A/A | 15.1 | 8.8 | 12.4 | 26.4 | 8.3 | 21.1 | 13.4% |
| Cytochrome P450 Family 1 Subfamily A Member 2 |  |  |  | ALL | AFR | AMR | EUR | EAS | SAS | SDC |
| CYP1A2 | Normal Metabolizer - Possible Inducibility | Ancestral: C or G | *1A/*1A (C/C or G/G), *1A/*1V, *1A/*1W | 15.2 | 19.8 | 7.5 | 10.9 | 11.5 | 22.7 | 23.7% |
| CYP1A2 | Normal Metabolizer - Higher Inducibility | rs762551 (-163)C>A | *1A/*1F (C/A), *1F/*1F (A/A) | 84.8 | 80.2 | 92.5 | 89.1 | 88.5 | 77.3 | 74.2% |
| CYP1A2 | Poor Metabolizer - Lower Inducibility | rs2069514 (-3860G > A) | *1A/*1C (G/A) | 28.9 | 44.5 | 38.9 | 4 | 40.3 | 14.7 | 1.0% |
| CYP1A2 | Unknown Phenotype | multiple (***) | *1L/*1L, *1L/*1W | N/A | N/A | N/A | N/A | N/A | N/A | 1.0% |
| Cytochrome P450 Family 2 Subfamily C Member 19 |  |  |  | ALL | AFR | AMR | EUR | EAS | SAS | SDC |
| CYP2C19 | Normal Metabolizer | Ancestral: G or C | *1/*1 (G/G or C/C) | 33.7 | 27.5 | 57.6 | 31.8 | 42.1 | 18.8 | 29.5% |
| CYP2C19 | Intermediate Metabolizer | rs4244285 (19154G>A) | *1/*2 (G/A) | 32.8 | 27.1 | 19.3 | 26.6 | 47.4 | 41.3 | 34.4% |
| CYP2C19 | Poor Metabolizer | rs4244285 (19154G>A) | *2/*2 (A/A) | 5.8 | 3.5 | 0.9 | 1.2 | 7.5 | 15.1 | 15.6% |
| CYP2C19 | Rapid Metabolizer | rs12248560 (-806C>T) | *1/*17 (C/T) | 24.7 | 36.8 | 20.5 | 36 | 3 | 22.3 | 20.5% |
| CYP2C19 | Ultra-Rapid Metabolizer | rs12248560 (-806C>T) | *17/*17 (T/T) | 3 | 5.1 | 1.7 | 4.4 | 0 | 2.5 | 1.2% |
| Cytochrome P450 Family 2 Subfamily C Member 9 |  |  |  | ALL | AFR | AMR | EUR | EAS | SAS | SDC |
| *CYP2C9* | Normal Metabolizer | Ancestral: G or A | *1/*1 | 90.5 | 99.5 | 92.5 | 85.7 | 93.5 | 78.7 | 77.0% |
| *CYP2C9* | Intermediate Metabolizer | rs1057910 (A/C) | *1/*3 (A/C) | 9.3 | 0.5 | 7.5 | 14.1 | 6.3 | 20.7 | 19.7% |
| *CYP2C9* | Poor Metabolizer | rs1057910 (C/C) | *3/*3 (C/C) | 0.2 | 0 | 0 | 0.2 | 0.2 | 0.6 | 3.3% |
| **Cytochrome P450 Family 2 Subfamily D Member 6** |  |  |  | **ALL** | **AFR** | **AMR** | **EUR** | **EAS** | **SAS** | **SDC** |
| *CYP2D6* | Normal Metabolizer | Ancestral: multiple | *1/*1, *1/*2, *1/*4, *1/*5... | 71.5 | 82.8 | 82.3 | 88.0 | 41.5 | 79.6 | 89.7% |
| *CYP2D6* | Intermediate Metabolizer | *10 - rs1065852 (100C>T) | *5/*10, *10/*15, *4/*17, *4/*29, *4/*41 | 23.8 | 11.3 | 14.8 | 5 | 57.1 | 16.5 | 4.1% |
| *CYP2D6* | Poor Metabolizer | *4 - rs3892097 (1846G>A) | *4/*4, *4/*5 (A/A) | 2.1 | 1.2 | 2.9 | 4.6 | 0 | 2.5 | 3.7% |
| *CYP2D6* | Ultra-Rapid Metabolizer (****) | XN (Duplication, XN Exon 9) | *1/*2 XN, *1/*4 XN, *1/*35 XN | 2.64 | 4.66 | N/A | 2.37 | 1.37 | 1.37 | 2.5% |
| **Cytochrome P450 Family 3 Subfamily A Member 4** |  |  |  | **ALL** | **AFR** | **AMR** | **EUR** | **EAS** | **SAS** | **SDC** |
| *CYP3A4* | Normal Metabolizer | Ancestral: G | *1/*1 (G/G) | 97 | 99.8 | 94.8 | 90.3 | 100 | 98.8 | 67.6% |
| *CYP3A4* | Intermediate Metabolizer | rs35599367 (intron 6 C>T) | *1/*22 (G/A) | 3 | 0.2 | 5.2 | 9.7 | 0 | 1.2 | 28.4% |
| **Cytochrome P450 Family 3 Subfamily A Member 5** |  |  |  | **ALL** | **AFR** | **AMR** | **EUR** | **EAS** | **SAS** | **SDC** |
| *CYP3A5* | Normal Metabolizer | Ancestral: T | *1/*1 (T/T) | 22.7 | 67.6 | 5.8 | 0.4 | 7.9 | 12.1 | 5.3% |
| *CYP3A5* | Intermediate Metabolizer | rs776746 (6986A>G) | *1/*3 (T/C) | 30.3 | 28.7 | 29.1 | 10.5 | 41.5 | 42.3 | 45.8% |
| *CYP3A5* | Poor Metabolizer | rs776746 (6986A>G) | *3/*3 (C/C) | 47 | 3.6 | 65.1 | 89.1 | 50.6 | 45.6 | 54.2% |
| **Dopamine Receptor D2** |  |  |  | **ALL** | **AFR** | **AMR** | **EUR** | **EAS** | **SAS** | **SDC** |
| *DRD2* | Homozygous for rs1799978 C allele | Ancestral: C | C/C | 2.2 | 4.1 | 0.3 | 0.6 | 1 | 0.8 | 1.3% |
| *DRD2* | Heterozygous for rs1799978 C allele | rs1799978 (-241A>G) | T/C | 19.5 | 26.5 | 14.7 | 10.7 | 28 | 13.7 | 12.3% |
| *DRD2* | Homozygous for rs1799978 T allele | rs1799978 (-241A>G) | T/T | 78.4 | 69.4 | 85 | 88.7 | 68.3 | 85.5 | 86.4% |
|  |  |  |  | **ALL** | **AFR** | **AMR** | **EUR** | **EAS** | **SAS** | **SDC** |
| *DRD2* | Homozygous for rs2283265 C allele | Ancestral: C | C/C | 62 | 85 | 55.9 | 73.6 | 34.3 | 51.9 | 41.6% |
| *DRD2* | Heterozygous for rs2283265 A allele | rs2283265 (724-353G>T) | C/A | 30.6 | 13.8 | 35.2 | 23.3 | 48.4 | 39.1 | 32.4% |
| *DRD2* | Homozygous for rs2283265 A allele | rs2283265 (724-353G>T) | A/A | 7.4 | 1.2 | 8.9 | 3.2 | 17.3 | 9 | 26.1% |
| **5-Hydroxytryptamine Receptor 2A (Serotonin 2A receptor gene)** |  |  |  | **ALL** | **AFR** | **AMR** | **EUR** | **EAS** | **SAS** | **SDC** |
| *HTR2A* | Homozygous for G allele (rs7997012) | Ancestral: G | G/G | 56.2 | 97.1 | 41.8 | 33.2 | 56.7 | 34.2 | 7.2% |
| *HTR2A* | Heterozygous for the A allele (rs7997012) | rs7997012 (614-2211T>C) | A/G | 33.1 | 2.9 | 46.1 | 47.7 | 34.9 | 47.6 | 30.8% |
| *HTR2A* | Homozygous for the A allele (rs7997012) | rs7997012 (614-2211T>C) | A/A | 10.7 | 0 | 12.1 | 19.1 | 8.3 | 18.2 | 62.0% |
|  |  |  |  | **ALL** | **AFR** | **AMR** | **EUR** | **EAS** | **SAS** | **SDC** |
| *HTR2A* | Homozygous for the C allele (rs6311) | Ancestral: C | C/C | 32.4 | 36 | 40.1 | 33 | 18.3 | 36 | 25.2% |
| *HTR2A* | Heterozygous for the T Allele (rs6311) | rs6311 (-1438G>A) | C/T | 46.5 | 46.1 | 47.6 | 46.5 | 45.8 | 47 | 63.8% |
| *HTR2A* | Homozygous for the T allele (rs6311) | rs6311 (-1438G>A) | T/T | 21.1 | 17.9 | 12.4 | 20.5 | 35.9 | 17 | 11.0% |
| **5-Hydroxytryptamine Receptor 2C (Serotonin 2C receptor gene)** |  |  |  | **ALL** | **AFR** | **AMR** | **EUR** | **EAS** | **SAS** | **SDC** |
| *HTR2C* | Homozygous for the C allele (rs1414334) | Ancestral: C | C/C | 35 | 15.7 | 41.2 | 39 | 50.2 | 40.3 | 52.5% |
| *HTR2C* | Heterozygous for the C allele (rs1414334) | rs1414334 (2565G>C or 114138144C>G) | G/C | 51.9 | 49.4 | 53.4 | 51.3 | 49.2 | 54.4 | 41.9% |
| *HTR2C* | Homozygous for the G allele (rs1414334) | rs1414334 (2565G>C or 114138144C>G) | G/G | 13.1 | 34.9 | 5.4 | 9.7 | 0.6 | 5.3 | 5.5% |
|  |  |  |  | **ALL** | **AFR** | **AMR** | **EUR** | **EAS** | **SAS** | **SDC** |
| *HTR2C* | Homozygous for the C allele (rs3813929) | Ancestral: C | C/C | 88.1 | 98.2 | 82.1 | 85.5 | 85.5 | 78.1 | 85.5% |
| *HTR2C* | Heterozygous for the C allele (rs3813929) | rs3813929 (-759C>T) | T/C | 10.7 | 1.8 | 17.9 | 13.5 | 13.5 | 17.6 | 12.8% |
| *HTR2C* | Homozygous for the T allele (rs3813929) | rs3813929 (-759C>T) | T/T | 1.2 | 0 | 0 | 1 | 1 | 4.3 | 1.7% |
| **Opioid Receptor Mu 1** |  |  |  | **ALL** | **AFR** | **AMR** | **EUR** | **EAS** | **SAS** | **SDC** |
| *OPRM1* | Normal OPRM1 Function | Ancestral: A | A/A | 62.5 | 98.2 | 64.6 | 70.2 | 36.7 | 31.3 | 53.0% |
| *OPRM1* | Altered OPRM1 Function | rs1799971 (A118G) | A/G | 30.4 | 1.8 | 30.8 | 27.2 | 48 | 53.8 | 47.0% |
| *OPRM1* | Altered OPRM1 Function | rs1799971 (A118G) | G/G | 7.1 | 0 | 4.6 | 2.6 | 15.3 | 14.9 | 0.0% |
| **Solute Carrier Family 6 Member 4** |  |  |  | **ALL** | **AFR** | **AMR** | **EUR** | **EAS** | **SAS** | **SDC** |
| *SLC6A4* | Homozygous for C Allele | Ancestral: C | C/C | 28.7 | 3.8 | 30.5 | 17.9 | 68.4 | 31 | 16.3% |
| *SLC6A4* | Heterozygous for the C Allele | rs1042173 (463T>G C/A) | C/A | 39.7 | 29.5 | 47.3 | 51.7 | 27.6 | 48.3 | 64.8% |
| *SLC6A4* | Homozygous for A Allele | rs1042173 (463T>G C/A) | A/A | 31.6 | 66.7 | 22.2 | 30.4 | 4 | 20.7 | 18.9% |
| *SLC6A4* | Normal Serotonin Transporter Expression | 5-HTTLPR (L/S) and rs25531 (A/G) | La/La (L'L' group**) | N/A | 27 | 22 | 25 | 8 | 8 | 24.7% |
| *SLC6A4* | Decreased Serotonin Transporter Expression | 5-HTTLPR (L/S) and rs25531 (A/G) | La/Lg, La/S (L'S' group**) | N/A | 49 | 51 | 50 | 30 | 30 | 43.8% |
| *SLC6A4* | Low Serotonin Transporter Expression | 5-HTTLPR (L/S) and rs25531 (A/G) | Lg/Lg, Lg/S, S/S (S'S' group**) | N/A | 24 | 27 | 25 | 62 | 62 | 31.5% |
| **Solute Carrier Organic Anion Transporter Family Member 1B1** |  |  |  | **ALL** | **AFR** | **AMR** | **EUR** | **EAS** | **SAS** | **SDC** |
| *SLCO1B1* | Normal Function | Ancestral: T | T/T | 43.5 | 72.9 | 27.4 | 15.1 | 57.1 | 30.3 | 81.6% |
| *SLCO1B1* | Decreased Function | rs4149057 (521T>C) | T/C | 39.5 | 25.3 | 44.4 | 48.3 | 37.1 | 48.9 | 16.8% |
| *SLCO1B1* | Poor Function | rs4149057 (521T>C) | C/C | 17 | 1.8 | 28.2 | 36.6 | 5.8 | 20.8 | 1.6% |
| **Vitamin K Epoxide Reductase Complex Subunit 1** |  |  |  | **ALL** | **AFR** | **AMR** | **EUR** | **EAS** | **SAS** | **SDC** |
| *VKORC1* | Low Warfarin Sensitivity | Ancestral: G | G/G | 50.9 | 89.5 | 35.2 | 38.2 | 1.8 | 73.4 | 49.8% |
| *VKORC1* | Intermediate Warfarin Sensitivity | rs9923231 (-1639G>A) | G/A | 27.1 | 10 | 47.6 | 46.1 | 19.4 | 24.1 | 39.7% |
| *VKORC1* | High Warfarin Sensitivity | rs9923231 (-1639G>A) | A/A | 22 | 5 | 17.2 | 15.7 | 78.8 | 2.5 | 10.5% |

\* The frequencies for this table were referenced from the 1000 Genomes Database Ensembl which is available at http://grch37.ensembl.org/Homo_sapiens/Info/Index. Further information is available at http://www.cypalleles.ki.se/. Populations have been divided into 5 super populations: African (**AFR**), South Asian (**SAS**), Ad Mixed American (**AMR**), East Asian (**EAS**), European (**EUR**), this study (San Diego Cohort: **SDC**)
\*\* Group definition as per Pascale E, Ferraguti G, Codazzo C, Passarelli F, Mancinelli R, Bonvicini C, et al. Alcohol Dependence and Serotonin Transporter Functional Polymorphisms 5-HTTLPR and rs25531 in an Italian Population. 2015;1–7. Population frequencies for *SLC6A4* 5-HTTLPR (L/S), rs25531 (A/G) derived from Haberstick BC, Smolen A, Williams RB, Bishop GD, Foshee VA, Thornberry TP, et al. Population frequencies of the Triallelic 5HTTLPR in six Ethnically diverse samples from North America, Southeast Asia, and Africa. Behav Genet. NIH Public Access; :45:255–61. Available from: http://www.ncbi.nlm.nih.gov/pubmed/25564228
\*\*\* See Soyama A, Saito Y, Hanioka N, Maekawa K, Komamura K, Kamakura S, et al. Single nucleotide polymorphisms and haplotypes of *CYP1A2* in a Japanese population. Drug Metab Pharmacokinet. 2005;20:24–33.
\*\*\*\* Based on Beoris M, Wilson JA, Garces JA, Lukowiak AA. CYP2D6 copy number distribution in the US population. Pharmacogenet Genomics. 2016;26:96–9.

Of the 171 patients studied, drug adherence data was not available for 69 patients for which PGx report data was summarized. PGx report implementation could only be studied on the remaining 102 patients. 26 PGx reports showed no medication list provided by the clinic, 8 of which medication lists were made available and added onto the PGx report retroactively. Medication lists provided showed that patients were prescribed an average of 5 different medications (ranging from 0-25 medications), resulting on average in 1 moderate pharmacogenetic guidance and 3 moderate drug-drug-interaction observations per patient. Among patient PGx reports with medication lists provided (n = 146) 57.5% showed one or more moderate and 5.5% at least one serious pharmacogenetic interaction. 66% of patients showed at least one moderate and 15% one or more serious drug-gene or drug-drug-interaction (**Fig 3, Supplementary Table 5**).

**Fig 3.**
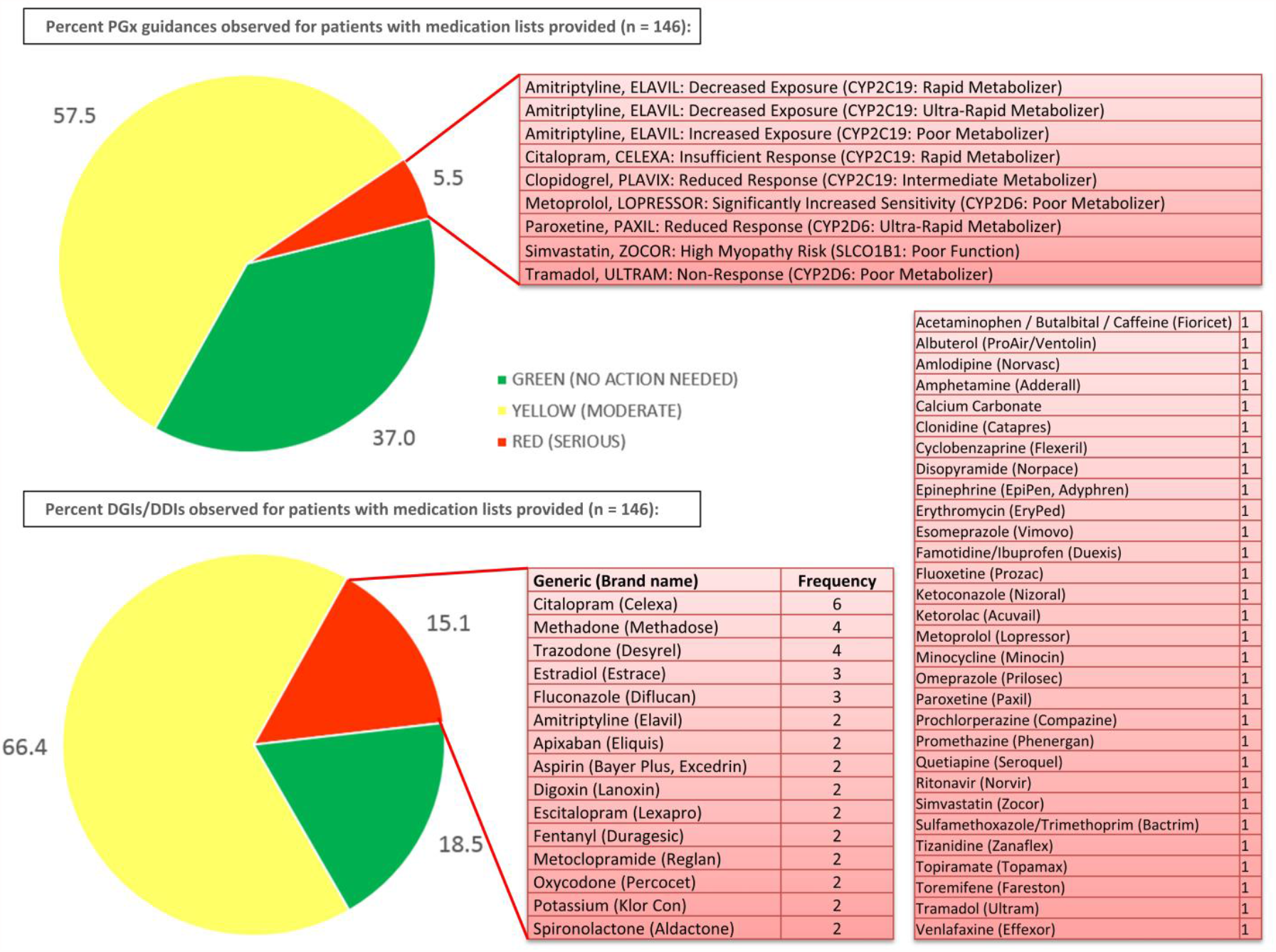
Percent genetic (PGx) dosing guidance, DGI (drug-gene-interactions) and DDI (drug-drug-interactions) observed for patients with medication lists provided (n = 146) sorted by expected normal response to a drug based on PGx, or no interaction observed for DGIs/DDIs. GREEN – no action required, YELLOW (moderate) or RED (serious) interactions prompting actionable PGx or DGI/DDI recommendations. Specific drug names and the associated genotype for PGx dosing or frequency for DGIs/DDIs are shown for serious cases.

Medications affecting patients most severely based on their individual genotype in this cohort were amitriptyline for decreased exposure among 2 *CYP2C19* rapid metabolizers and increased exposure for 1 *CYP2C19* poor metabolizer, citalopram (insufficient response, *CYP2C19* rapid metabolizer), clopidogrel (reduced response, *CYP2C19* intermediate metabolizer), metoprolol with significantly increased sensitivity for a *CYP2D6* poor metabolizer, paroxetine (reduced response in *CYP2D6* ultra-rapid metabolizer), simvastatin (poor function of *SLCO1B1* inducing high myopathy risk) and tramadol (*CYP2D6* poor metabolizer with risk for no response). The top 15 medications affecting patients based on a DGI or DDI were identified (**Fig 3**). The most frequently occurring moderate drug-drug-interaction involved opioids observed in combination with CNS depressants such as muscle relaxants, benzodiazepines, sleep drugs or the nerve pain medications gabapentin and pregabalin (**Supplementary Table 5**).

Prescription regimens were determined for 102 patients based on drug adherence report data before and after the PGx report was made available. Remaining patients either showed no drug adherence data or limited drug adherence data before the PGx report but no further information afterwards. An active change in prescriptions based on the PGx report was observed for 85 patients (83%) for which a specific drug was either discontinued, switched within the defined drug classes of the report or a new drug added. 17% of patient reports showed no predictive evidence of ADRs even when prescribed up to 11 medications (on average 2.5 medications per patient). Appropriately, no action was taken by the provider in these cases to deviate from the original prescription regimen. All adjustments made to patient prescriptions were studied for potential contraindications or possible new ADRs based on the PGx report.

Of the 102 patients whose medication lists were adjusted only 3 showed recommendations in the PGx report were not being followed for unknown reasons. “Patient A” was shown to be administered 5 medications (KEFLEX®, PENNSAID®, SKELAXIN®, MS CONTIN® and LIDOCAINE CV®). PGx reporting indicated a normal PGx response and one moderate DDI to MS CONTIN® (morphine) and SKELAXIN® (metaxalone) and a moderate PGx interaction for PENNSAID® (diclofenac). Cessation of SKELAXIN® and PENNSAID® removed all moderate ADRs, however, addition of PERCOCET® (oxycodone and acetaminophen) was not recommended: “*Oxycodone - CYP2D6 Poor Metabolizer. Test results indicate a possible increased risk of therapeutic failure. Monitor for decreased response or may select alternative medication*.” The decreased response was alleviated with morphine prescriptions, for which there were no contraindications. Progress notes showed Patient A “*has tried to use topical patches but experienced a localized reaction to the adhesive on the patch. Oral pain medication of MS Contin and Percocet is helpful*. Patient A *notes that some days* Patient A *does not require the max dose of the Percocet*.” COREG® (carvedilol) was added to prescription regimen causing a moderate PGx warning: “*CYP2D6 Poor Metabolizer: Test results indicate an increased risk of dizziness during up-titration. Consider standard prescribing and monitoring practices with careful dose titration*.” The addition of SILENOR® (doxepin) was also contraindicated by the PGx report: “*CYP2D6 Poor Metabolizer: Test results indicate an increased risk of adverse effects. Consider an alternative medication or a 50% dose reduction with therapeutic drug monitoring*.” In this case the prescribed doxepin dosage was minimal (10 mg/day) according to progress notes. For the treatment of major depression or anxiety adult oral dosages are initially 75 mg/day. Addition of WELLBUTRIN® (bupropion), SOMA® (carisoprodol), TOPAMAX® (topiramate) and PRILOSEC® (omeprazole) showed no contraindication except a moderate DDI between carisoprodol and morphine. The dose reduction for doxepin and remaining moderate interaction for carvedilol were acceptable as carvedilol was discontinued and appropriate monitoring practices were carried out for patient A.

Similarly, for “Patient B” 7 medications were listed, which showed a switch from codeine to morphine although no warnings against codeine were indicated (patient *CYP2D6* normal metabolizer status). Instead, a switch to morphine warned: *“The patient does not carry the COMT Val158Met variant. The patient may require higher doses of morphine for adequate pain control*”. Additionally, quetiapine and citalopram could cause a serious DDI (“*concurrent use with agents known to prolong the QT interval should be avoided*”), as well as the combinations of opioids with gabapentin prompted to “*monitor patients for gabapentinoid-related side effects*.” Further investigation into progress notes for Patient B showed a suspected allergy or adverse drug reaction to hydrocodone and oxycodone resulting in “nausea”, possibly explaining the emphasis on morphine and the patient avoiding exposure to other opioids such as codeine, hydrocodone or oxycodone. An increase in morphine 15 mg immediate release formulation tablets (MSIR®) was initiated from 3 to 4 daily, eventually 15 mg MSIR® 3x/day with an additional 15 mg MS CONTIN® (extended release) 2x/day: Patient B “*has tried and failed following medications: anti-inflammatory meds, hydrocodone and oxycodone/oxycontin in the past. Patient reports the medication initiated last office visit has provided better relief in pain, notes oral pain medications in form of MSIR and MS Contin are effective and decreases low back pain by no less than a 60% relief in pain, pain level today is 6/10. Upon questioning patient denies adverse reactions such as euphoria/dysphoria*”. Monthly reviews of the patient’s condition show “*Denies trouble breathing, shortness of breath, asthma, sleep apnea, seizures, blackouts, trouble with memory, headache, fainting spells, numbness, weakness and tremors*.”

Patient C was maintained on 10 of 11 initial medications with the appropriate removal of PLAVIX® (clopidogrel) after 2 serious PGx warnings: “*Reduced Response to Clopidogrel (CYP2C19: Intermediate Metabolizer) Consider alternative therapy*” and “*High Myopathy Risk (SLCO1B1: Poor Function). Simvastatin plasma concentrations are expected to be elevated. Consider avoiding simvastatin and prescribe an alternative statin, or consider prescribing simvastatin at a lower starting dose (20 mg/day). Routine creatine kinase (CK) monitoring is also advised. The FDA recommends against the 80 mg daily dose*.” An additional serious DDI for ZOCOR® (simvastatin) and NORVASC® (amlodipine) warned “*do not exceed a dosage of 20 mg daily of simvastatin in patients receiving concurrent therapy with amlodipine. If concurrent therapy is deemed medically necessary, monitor patients for signs and symptoms of myopathy/rhabdomyolysis, including muscle pain/tenderness/weakness, fever, unusual tiredness, changes in the amount of urine and/or discolored urine*.” After PGx reporting, clopidogrel was no longer observed in medication lists for drug adherence reports, but simvastatin was continued with amlodipine and 9 moderate DDIs remained cautioning to “*limit the dosages and duration of each drug to the minimum possible while achieving the desired clinical effect*”. The only alternative statin without adverse interactions recommended was fluvastatin. Progress notes for Patient C showed simvastatin was prescribed less than 80 mg/day as recommended by the FDA in the PGx report at 40 mg/day. Patient C “*Denies muscle cramp, muscle twitches, muscle wasting, muscle weakness, neck pain, joint swelling. Denies fever, fatigue*”, however Patient C eventually reported “*muscle pain or tenderness*” in the latter of the 2-year treatment window. Monthly urinalysis screens and blood testing showed no discoloration in urine or abnormal glomerular filtration rates, but the reported muscle pain/tenderness and the combination of reduced *SLCO1B1* gene function with concurrent daily 40 mg simvastatin and 5 mg amlodipine possibly indicated a statin-induced myopathy [23].

## 4 Conclusion

In summary, the effect of PGx reports made available to the medical staff in this context seems quite significant as observed by the individual PGx, DGI and DDI recommendations showing a corresponding modification of the medication regimen for each patient. Preventative action was observed for all serious interactions and only moderate interactions were tolerated where there may not have been other alternatives. This study demonstrates the predictive value of PGx testing combined with a customized informational report to help improve clinical outcomes, which resulted in increased utilization on patients in a pain management setting. While PGx cannot predict all adverse drug reactions (for example, allergies cannot be detected), dosing guidance and the additional drug-gene and drug-drug-interaction algorithm provided valuable insight to optimize prescription regimens.

## Data Availability

This study was conducted in accordance with the Declaration of Helsinki with written informed consent from each patient. Patient data collection and summaries were approved by the Alcala Pharmaceutical Inc. Institutional Review Board (IORG0010127, IRB00012026, #R003). All test samples derived from human subjects were de-identified of their health information as defined by HIPAA guidelines. Patient data for comparison of urine drug adherence testing before and after PGx reporting were obtained retrospectively from patients (n = 171) during routine testing in a pain management clinic representing an ethnically diverse patient population served by Alcala Testing and Analysis Services (ATAS) from 2016 to 2018 within the western United States. Available data includes de-identified pre- and post-PGx medication lists, PGx and urine drug adherence data.

## Author Contributions

Conceptualization, C.T. and D.J.S; methodology, C.T.; validation, C.T., M.J.C.M; formal analysis, M.J.C.M., C.T.; Resources, D.J.S.; writing - original draft preparation, C.T., writing - review and editing, C.T., M.J.C.M; supervision, D.J.S.; project administration, C.T.

## Employment or Leadership

C.T. and M.J.C.M are current employees of Alcala Testing and Analysis Services (ATAS). David J. Smith is a stakeholder of Alcala Testing and Analysis Services (ATAS).

## Acknowledgments

None declared.

## Testimony

None declared.

## Patents

None declared.

